# Epigenetic Alteration of Smooth Muscle Cells Regulates Endothelin-Dependent Blood Pressure and Hypertensive Arterial Remodeling

**DOI:** 10.1101/2024.07.09.24310178

**Authors:** Kevin D. Mangum, Qinmengge Li, Tyler M. Bauer, Sonya J. Wolf, James Shadiow, Jadie Y. Moon, Emily C. Barrett, Amrita D. Joshi, Zara Ahmed, Rachael Wasikowski, Kylie Boyer, Andrea T. Obi, Frank M. Davis, Lin Chang, Lam C. Tsoi, Johann Gudjonsson, Katherine A. Gallagher

**Affiliations:** Section of Vascular Surgery, Department of Surgery, University of Michigan, Ann Arbor, MI; Department of Microbiology and Immunology, University of Michigan, Ann Arbor, MI; Department of Internal Medicine, University of Michigan, Ann Arbor, MI

## Abstract

Long-standing hypertension (HTN) affects multiple organ systems and leads to pathologic arterial remodeling, which is driven largely by smooth muscle cell (SMC) plasticity. Although genome wide association studies (GWAS) have identified numerous variants associated with changes in blood pressure in humans, only a small percentage of these variants actually cause HTN. In order to identify relevant genes important in SMC function in HTN, we screened three separate human GWAS and Mendelian randomization studies to identify SNPs located within non-coding gene regions, focusing on genes encoding epigenetic enzymes, as these have been recently identified to control SMC fate in cardiovascular disease. We identified SNPs rs62059712 and rs74480102 in the promoter of the human *JMJD3* gene and show that the minor C allele increases *JMJD3* transcription in SMCs via increased SP1 binding to the *JMJD3* promoter. Using our novel SMC-specific Jmjd3-deficient murine model (*Jmjd3^flox/flox^Myh11^CreERT^*), we show that loss of *Jmjd3* in SMCs results in HTN, mechanistically, due to decreased *EDNRB* expression and a compensatory increase in *EDNRA* expression. As a translational corollary, through single cell RNA-sequencing (scRNA-seq) of human arteries, we found strong correlation between *JMJD3* and *EDNRB* expression in SMCs. Further, we identified that JMJD3 is required for SMC-specific gene expression, and loss of JMJD3 in SMCs in the setting of HTN results in increased arterial remodeling by promoting the SMC synthetic phenotype. Our findings link a HTN-associated human DNA variant with regulation of SMC plasticity, revealing therapeutic targets that may be used in the screening and/or personalized treatment of HTN.

**Graphical Abstract:** 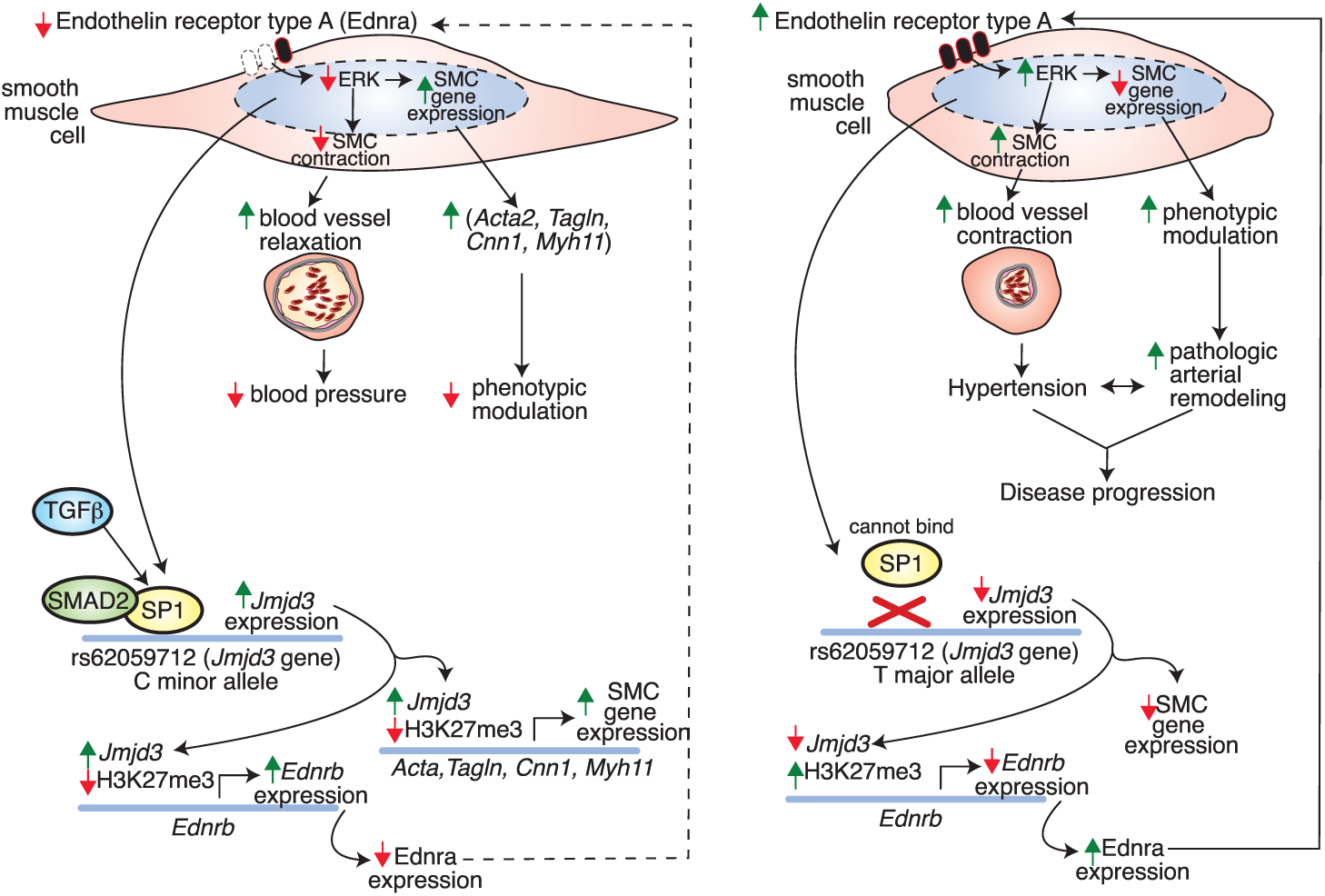

## Introduction

Hypertension (HTN) contributes to significant morbidity and mortality in the United States primarily due to its detrimental effects on end-organs including the heart and blood vessels, as well as others [1]. Elevated blood pressure is directly regulated by peripheral vascular resistance, which is controlled by blood vessel tone, a process mediated by vascular smooth muscle cells (SMCs) [2]. Proper regulation of blood pressure requires appropriate vascular SMC contraction or relaxation, and is largely dependent on SMC phenotype. SMC function is regulated by numerous pharmacologic and mechanical stimuli. In this regard, endothelin-1, a potent vasoconstrictor, binds to the endothelin receptor, leading to myosin light chain phosphorylation and SMC contraction. Two endothelin receptors are expressed in SMCs (A and B), however, EDNRA is considered the predominant receptor that mediates SMC contractility and vessel tone [3]. In contrast, the exact role of EDNRB in mediating SMC contraction versus relaxation is less established, since recent reports have indicated dual roles for EDNRB [3–5]. Endothelin receptor activation is a key pathway for SMC contractility and HTN, and endothelin receptor blockade decreases blood pressure in experimental models of HTN [4]. SMC contractility/function is largely dictated by SMC phenotype, which is highly plastic with the ability to switch between a contractile or synthetic phenotype, depending on upstream cues. The contractile phenotype is induced by many factors, including mechanical (e.g., cell stretch) and molecular (e.g., TGFβ) stimuli [6, 7]. SMC differentiation to the contractile phenotype is regulated by the transcription factors, serum response factor (SRF), myocardin, and the myocardin-related transcription factors (MRTFs) [8–11]. Other transcription factors such as SP1 and the SMAD family induce genes required for the contractile SMC phenotype [12, 13]. In contrast, SMC transition to the synthetic phenotype occurs in diseases including atherosclerosis, hypertension, and restenosis and is driven by factors including PDGF, inflammatory cytokines, and BMP [6, 14, 15]. The transcription factors, KLF4 and ELK1, repress SMC differentiation and are associated with the synthetic phenotype [16, 17]. The precise regulation of SMC differentiation by these upstream cues is highly relevant in cardiovascular disease during which vascular SMCs become phenotypically modulated and contribute to pathology. In these pathologic processes, SMC genes (*ACTA2, TAGLN, CNN1, MYH11*) are repressed while there is an upregulation of proliferative-associated genes (*KLF4, FOS*) [6, 18]. Although the upstream signaling pathways have been explored, many of the underlying molecular and epigenetic mechanisms that regulate SMC phenotypes in cardiovascular pathologies, particularly HTN, remain unknown. Epigenetic mechanisms have been previously shown by our group and others to regulate cell phenotypes and control downstream gene expression in homeostatic and pathologic states [19–21]. In particular, epigenetic alteration of chromatin structure by epigenetic enzymes and key transcription factors has been shown to influence SMC phenotype during development and cardiovascular pathology [22–28]. Given that epigenetic processes have been partly explored in SMCs in the setting of vascular disease, identification of the precise epigenetic mechanisms regulating SMC phenotype are necessary to develop targeted therapies for cardiovascular diseases.

Genome wide association studies (GWAS) and similar large-scale approaches (e.g., Mendelian randomization) have recently identified numerous genetic loci that influence cardiovascular disease and can thus be used to prioritize novel candidate genes encoding epigenetic enzymes that may play a role in human disease [29]. In this regard, one recent GWAS for blood pressure in humans identified the SNP rs62059712 (major allele), which was associated with increased systolic blood pressure (SBP) [30]. Additionally, a separate GWAS for medication use associated rs62059712 with anti-hypertensive agents targeting the renin-angiotensin system [31]. Further, a Mendelian randomization study linked the rs62059712 major allele with increased SBP and pulse pressure in humans [32]. According to the HaploReg v4 database, the BP-associated allele in the European population is currently defined by two single nucleotide polymorphisms (SNPs rs62059712 and rs74480102) in high linkage disequilibrium (LD) (defined as r^2^>0.3 and D’=1) [33]. Notably, these two SNPs are located upstream of an epigenetic regulator, the histone demethylase, *JMJD3*. This is relevant, given JMJD3 has been previously studied by our group and others, where it plays a role in abdominal aortic aneurysm formation and controls inflammatory gene expression in innate immune cells in cardiometabolic disease [34, 35]. Given that three separate large human studies identified a significant variant in the *JMJD3* promoter that could influence HTN as well as JMJD3’s previously identified relevance in cardiovascular disease, we examined if this human data is relevant for SMC function and HTN. Specifically, whether the major allele variant within the SBP-locus regulates *JMJD3* gene transcription and if alterations in *JMJD3* in SMCs influence cell phenotype and cause pathologic changes in blood pressure remain unknown.

Here, we provide a mechanistic link between the human rs62059712 variant on *JMJD3* transcription in human and murine SMCs and the consequence of JMJD3 deficiency on blood pressure, SMC phenotype, and downstream arterial remodeling *in vivo*. SNP rs62059712 is located in the promoter of *JMJD3* (a histone demethylase), suggesting that rs62059712 or other SNPs in linkage with rs62059712 may affect *JMJD3* expression. We identified that the rs62059712 major T allele (mean allele frequency, MAF 0.92) was associated with increased blood pressure in humans. The minor C (protective) allele increased *JMJD3* transcription in SMCs by promoting SP1 transcription factor binding to the *JMJD3* promoter. SMC-specific deletion of *Jmjd3* in mice (*Jmjd3^flox/flox^Myh11^CreERT^*) resulted in increased blood pressure in an Angiotensin II model of HTN, which was due to decreased *EDNRB* and increased *EDNRA,* genes encoding expression of Endothelin Receptors A and B, respectively, in SMCs. Endothelin receptor antagonism eliminated increased vessel contractility after JMJD3 deletion. Additionally, JMJD3 loss in SMCs enhanced endothelin-ERK activation, resulting in increased SMC phenotypic modulation in response to vascular injury (i.e. chronic hypertension). Our study provides a mechanistic link between SNPs in the human population, *JMJD3* expression in SMCs associated with blood pressure alterations, and an SMC phenotypic switch in a murine model, and also identifies a previously unrecognized genetic target that may be used to develop novel therapies in patients with HTN.

## Methods

### Animals

Animal studies were approved by the Animal Care Committee of the University of Michigan and complied with the National Institutes of Health (NIH) guide for the care and use of laboratory animals. Mice were housed in stock-holding rooms under specific pathogen-free conditions. *Jmjd3^flox/flox^Myh11^CreERT^*and *Jmjd3^flox/flox^Tagln^Cre^* mice were bred on a C57BL6/J background, and experiments were performed when mice were 8-12 weeks old. For inducible deletion of *Jmjd3* in SMCs, mice were injected intraperitoneally with tamoxifen (75 mg/kg) for 5 consecutive days, then allowed a 3-day washout period before proceeding with experiments. Mice had free access to water and food throughout the study.

### Angiotensin II infusion and non-invasive blood pressure measurements

After 3 days of tamoxifen washout, osmotic mini-pumps (ALZET, Model 2004) containing saline or Angiotensin II were inserted subcutaneously in mice as previously described [34]. Angiotensin II was infused at a rate of 1 ug/kg/min. Mice were allowed to recover for 1 day before commencement of blood pressure measurements. Daily blood pressure measurements were obtained using noninvasive tail-cuff system (Kent Scientific). Animals were allowed to equilibrate in the system and at least two cycles of preliminary blood pressure measurements were performed to ensure accurate recording. Blood pressure was measured daily through day 14 at which time mice were sacrificed and tissues were harvested for downstream analysis.

### Aortic and mesenteric artery ring contractility assays

Aortas and mesenteric arteries were isolated after 5 days of intraperitoneal tamoxifen (75 mg/kg) or corn oil injection of *Jmjd3^flox/flox^Myh11^CreERT^*mice, following a 3-day washout period. For each mouse, vessels were cut into two rings and run in parallel. Aortic and mesenteric rings were mounted vertically by sliding them into two wire hooks. Aortic rings were immersed in 2 ml KH buffer. KH buffer contained the following (in mM): 118 NaCl, 4.8 KCl, 1.2 MgSO4, 1.2 KH2PO4, 25 NaHCO3, 11 glucose, and 2.5 CaCl2. The pH of the buffer was adjusted and maintained at 7.4 with 95% O2-5% CO2 at 37C. Aortic rings were equilibrated for 90 min with a resting tension of 1 g. After baseline tension was determined, vessels were precontracted with 60 mM KCl. Increasing concentrations of endothelin-1 (10^-10^-10^-7^ M), angiotensin II (10^-9^-10^-5^ M), and phenylephrine (10^-9^-10^-4^ M) were used. Concentration of bosentan used was 10^-8^ M. Values were normalized to percent contraction of KCl.

### Cell culture

Human aortic and bronchial smooth muscle cells were purchased from Lonza and maintained in Clonetics Smooth Muscle Growth Medium-2 (SMGM-2) supplemented with growth factors, 5% FBS, and antibiotics. Human SMCs were maintained and used for experiments between passages 2 and 14. Primary mouse aortic SMCs (mAoSMCs) were isolated from the thoracic aortas of 12-16 week old C57BL6 mice and *Jmjd3^flox/flox^Tagln^Cre^* mice and maintained and used for experiments through passage 14 in DMEM F12 supplemented with 10% FBS and 0.5% penicillin and streptomycin.

### Plasmids

pGL3-*JMJD3* DHS 1 and 2 major and minor allele reporter constructs were generated by amplifying approximately 600 bp gene regions encompassing each SNP from a human SMC genomic DNA template. DNA fragments were then cloned into pGL3 luciferarase basic vector (Addgene), and the correct sequence was verified by Sanger sequencing. Mutations were introduced using the QuickChange site-directed mutagenesis protocol. All mutations were verified by Sanger sequencing.

### Luciferase assays

mAoSMCs were seeded in 24-well plates at a density of 2.4 x 10^4^ cells/well. Cells were transfected the day after plating with 50 ng of plasmid per well. Luciferase activity was measured 48 h after transfection using the Steady-Glo Luciferase Kit (Promega) according to the manufacturer’s instructions. Raw luciferase values were normalized to the activity of the promoterless pGL3 empty vector.

### Gel contractility assays

mAoSMCs were trypsinized when 80% confluent and resuspended at 6×10^5^ cells/mL in DMEM F12 with 10% FBS and antibiotics. Cells were diluted in type 1 collagen with DMEM F12 with 10% FBS and 0.5% antibiotics to give 1 mg/mL collagen and 3×10^5^ cells/mL final concentration. 500 ul of gel and cell mixture was added to each well of a 24-well plate. Gels were allowed to polymerize at 37C for 1 hour. Once gels were polymerized, gels were freed from well edges of the cell culture plate and 500 ul of media (with or without pharmacologic agent) was added to each well. Collagen gels were incubated for 24 hours, and the area of each gel was measured over this time.

### Scratch assays

Cells were seeded in a 6-well culture dish and grown to at least 90% confluence. On the day of the experiment, cells were treated with 5 ug/ml mitomycin C for 2 hours. A vertical stratch was made in each well with a p200 pipette tip, the media was removed, washed once with PBS, and fresh media was added. Scratch area was measured from 0 hours to 18 hours.

### siRNA knockdowns

mAoSMCs were transfected with 40 nM siRNA targeted to *Jmjd3*, *Sp1*, *Ednrb*, or a non-targeting control (NTC) siRNA for 72 hours. RNAiMax (Invitrogen) was used as the transfection reagent. Depending on experiment, cells were either left in 10% FBS media, serum starved, or serum starved then treated with agonist or pharmacologic agent.

### RNA extraction and qRT-PCR

For cultured SMCs, cells were collected directly in Trizol, RNA was extracted, and converted to cDNA using superscript cDNA synthesis kit (Invitrogen). 20-50 ng cDNA was used in downstream quantitative real-time Taqman PCR and normalized using 18S. For tissues, samples were snap frozen, pulverized using mortar and pestle, and collected in trizol. Homogenized tissue was further digested into single cell suspension using 20G needle and syringe. Samples underwent RNA extraction and downstream qRT-PCR as above.

### Western blotting

Cultured SMCs were lysed in RIPA buffer plus protease and phosphatase inhibitors and 1 mM DTT. For tissues, samples were snap frozen, pulverized using mortar and pestle, and collected in RIPA buffer with protease and phosphatase inhibitors and 1 mM DTT as above. Lysates were normalized by total protein concentration, sample buffer was added, and then samples were boiled for 5 minutes. 10 to 50 ug of protein was loaded on an SDS-PAGE gel and run. Proteins were transferred to nitrocellulose or PVDF membranes and blocked in either 3% BSA or 5% non-fat dry milk. Blocked membranes were incubated with primary antibodies overnight at 4C. Membranes were washed and incubated with species-specific HRP-linked secondary antibodies, washed, and developed. For densitometry results, Western blots were analyzed using NIH ImageJ software.

### Affinity purification of transcription factors

Human aortic SMC nuclear lysates were prepared using the NUPER nuclear extraction kit from ThermoScientific per manufacturer protocol. Nuclear extracts were diluted with two volumes of 1.3xPD buffer (1x: 10mM Hepes pH 7.4, 8% glycerol, 1mM MgCl2, 0.05% Tritonx100, DTT 1.3 mM, protease and phosphatase inhibitors). 5’ biotinylated sense and unmodified anti-sense 25 bp oligos (Sigma) were annealed, added to diluted pre-cleared nuclear lysates, and incubated with rotation for 10 min at room temperature. Streptavidin Dynabeads (Invitrogen) were added to the rotating mixture, incubated for another 30 min with rotation, and then beads were washed. Complexes were eluted in 2X sample buffer for analysis via SDS-PAGE and Western blotting.

### Immunoprecipitations

Cells were lysed as above for Western blotting. 5 ug of JMJD3 antibody was linked to Protein G Dynabeads, then linked antibody-beads were added to lysate and rotated overnight at 4C. Immunoprecipitated complexes were washed, eluted in sample buffer by boiling, run on SDS-PAGE gel, and analyzed via Western blotting.

### Chromatin Immunoprecipitation (ChIP) experiments

mAoSMCs were fixed for 10 minutes in 0.7% formaldehyde. The crosslinking reaction was quenched by incubating cells with 0.125 M glycine for 5 minutes. ChIP assays were performed using the Abcam ChIP kit (Abcam Cat# ab500) according to manufacturer’s instructions. Sheared chromatin was incubated overnight at 4C with 2 ug of JMJD3 antibody (Abcam), H3K27me3 antibody (Active Motif), or normal rabbit IgG antibody (Diagenode) as a negative control for non-specific binding. Eluted DNA was used in downstream qPCR assays.

### Histology and Immunofluorescence

Tissues were harvested from mice and fixed in 10% formalin for 24 hours then stored in 70% ethanol. Specimens were then embedded in paraffin and sectioned onto microscope slides. After deparaffinization, sections underwent hematoxylin and eosin staining or were processed for immunofluorescence. For immunofluorescent staining, tissue section underwent antigen retrieval in citric acid buffer (pH 6.0). Samples were then were permeabilized, blocked, and incubated with primary antibody (1:200) in blocking solution overnight at 4C in a humidity chamber. The following day, slides were washed in PBS, then incubated with fluorophore-conjugated secondary antibody (1:500) in PBS for 2 hours at room temperature. Slides were washed with PBS, mounted, allowed to dry overnight, and then imaged.

### RNA-seq experiments

Cultured SMCs from *Jmjd3^flox/flox^Tagln^Cre^*mice were plated in 6-well plates at approximately 60% confluence. Cells were harvested the day after initial plating. 3 biological replicates for each genotype were used. RNA isolation was performed using a RNeasy Kit (Qiagen) with DNAse digestion. Library construction and analysis of reads was performed as described previously [55]. Briefly, reads were trimmed using Trimmomatic and mapped using HiSAT2 [56, 57]. Read counts were performed using the feature-counts option from the subRead package followed by the elimination of low reads, normalization and differential gene expression using edgeR [58, 59]. Differential expression was performed on mapped reads using the taqwise dispersion algorithm in edgeR.

### scRNA-seq experiments

Generation of single-cell suspensions for scRNA-Seq was performed in the following manner as described by our group previously [35]. Briefly, following informed consent from patients and in accordance with University of Michigan IRB Study # HUM00098915, femoral artery specimens were harvested during femoral endarterectomy, femoral-femoral bypass, or aorto-bi-femoral bypass operations. Samples were digested overnight at 4C. Cells were strained and then combined in a 1:1 ratio for scRNA-Seq by the University of Michigan Advanced Genomics Core on the 10x Genomics Chromium System. Libraries were sequenced on the Illumina NovaSeq 6000 sequencer. NovaSeq was used as the sequencing platform to generate 151 bp paired-end reads. We conducted adapter trimming and quality control procedures as described previously [60] The reads were then mapped using STAR [61] to build human GRCh37, and gene expression levels were quantified and normalized by HTSeq [62] and DESeq2 [63], respectively. Negative binomial models in DESeq2 were used to conduct differential expression analysis. Data processing, including quality control, read alignment, and gene quantification, was conducted using the 10X Genomics Cell Ranger software. Seurat was then used for normalization, data integration, and clustering analysis [64]. All clustered cells were mapped to corresponding cell types by matching cell cluster gene signatures with putative cell type-specific markers.

### Statistics

GraphPad Prism software (RRID:SCR_002798) version 9.2.0 was used to analyze the data. All data were analyzed for normal distribution and then statistical significance between multiple groups was obtained using Student t-tests, ANOVA, or Pearson’s correlation where appropriate. All p-values less than or equal to 0.05 were considered significant.

### Study approval

All experiments using human samples were approved by the IRB at the University of Michigan (IRB #: HUM00098915) and were conducted in accordance with the principles in the Declaration of Helsinki.

## Results

### rs62059712 minor C allele increases JMJD3 transcription via enhanced SP1 binding to a regulatory region in the JMJD3 promoter

Alterations in SMC phenotype are hallmarks of cardiovascular diseases (e.g., atherosclerosis), where SMCs lose their mature, contractile markers, and switch to a synthetic, proliferative phenotype, which drives pathologic arterial remodeling [6]. Control of SMC phenotype is highly complex and context and disease-specific, and while several upstream regulators of SMC phenotype have been identified, there remains a knowledge gap in the specific downstream mechanisms that underlie SMC plasticity. Our group and others have identified that epigenetic pathways regulate cell plasticity in the context of cardiovascular disease [28, 36]. To identify translationally relevant epigenetic mechanisms in the regulation of blood pressure, we analyzed a large genome wide association study (GWAS) for blood pressure, a GWAS for anti-hypertensive medication use, and a Mendelian randomization study for hypertension (HTN) to identify human variants that were located within or near genes encoding various chromatin modifying enzymes (CMEs) that had been previously found to be relevant in cardiovascular disease [30–32]. Our most promising target was a SNP, rs62059712, that was located upstream of human *JMJD3*, a histone demethylase that has previously been shown to regulate macrophage phenotype in cardiometabolic diseases [34, 35, 37]. In these separate large studies for blood pressure, the rs62059712 major T allele was associated with increased blood pressure. In the GWAS study, the major allele was associated with a 0.39 mmHg increase in systolic blood pressure (SBP) per allele copy. Similarly, in the Mendelian randomization study, the major T allele was associated with a 0.46 mmHg increase in SBP and 0.32 mmHg increase in pulse pressure per allele copy. Furthermore, very interestingly, in the GWAS for anti-hypertensive medication use, the major T allele was associated with increased use of agents acting on the renin-angiotensin system (e.g., ACE inhibitors) [31]. Because these identified SNPs are not necessarily causative, we investigated whether rs62059712 or associated SNPs in linkage disequilibrium (LD) affected *JMJD3* transcription. We utilized the HaploReg web-based tool to identify SNPs in LD with rs62059712, and only rs74480102 reached LD threshold (defined in this study as r^2^>0.3).

To locate rs62059712 and rs74480102 in relation to the *JMJD3* gene and possible alignment with features of open chromatin, we utilized the publicly accessible UCSC Genome Browser [38]. Notably, rs62059712 and rs74480102 were both located upstream of the *JMJD3* gene within regions that aligned with several features of active transcription including H3K27 acetylation ChIP-seq peaks, DNase Hypersensitive Sites (DHS), and vertebral conservation **(Figure 1A)**. Thus, we hypothesized that rs62059712 (within DHS1) and/or rs74480102 (within DHS2) altered *JMJD3* transcription by influencing chromatin and transcription factor accessibility to one or both of these regions. Since SMC contractility directly regulates vascular tone, and thus blood pressure, we hypothesized that *JMJD3* expression in SMCs was important for blood pressure regulation. To test this, we generated an allelic series of luciferase reporter constructs corresponding to the major and minor alleles for both SNPs, transfected each into human SMCs (HuSMCs) and measured downstream luciferase activity. As shown in **Figure 1B**, in luciferase assays, the rs62059712 minor C allele demonstrated increased transcriptional activity compared to the major T allele in HuSMCs. Notably, there was no difference in luciferase activity between the rs74480102 major G and minor A alleles **(Figure 1C)**. Further, the rs62059712 minor C allele exhibited increased luciferase activity compared to the major T allele in mouse aortic SMCs (mAoSMCs), and there was no difference in transcription activity between rs74480102 major or minor alleles **(Supplemental Figure 1 A-B)**. Next, to test the effect of the rs62059712 allele variant on *JMJD3* gene expression in human SMCs, we used CRISPR-Cas9 technology to delete the 450 bp region encompassing the SNP (DHS1) in HuSMCs. Deletion of the 450 bp DHS1 region resulted in decreased *JMJD3* expression, further supporting the role of rs62059712 in regulating *JMJD3* expression **(Figure 1D)**.

**Figure 1.**
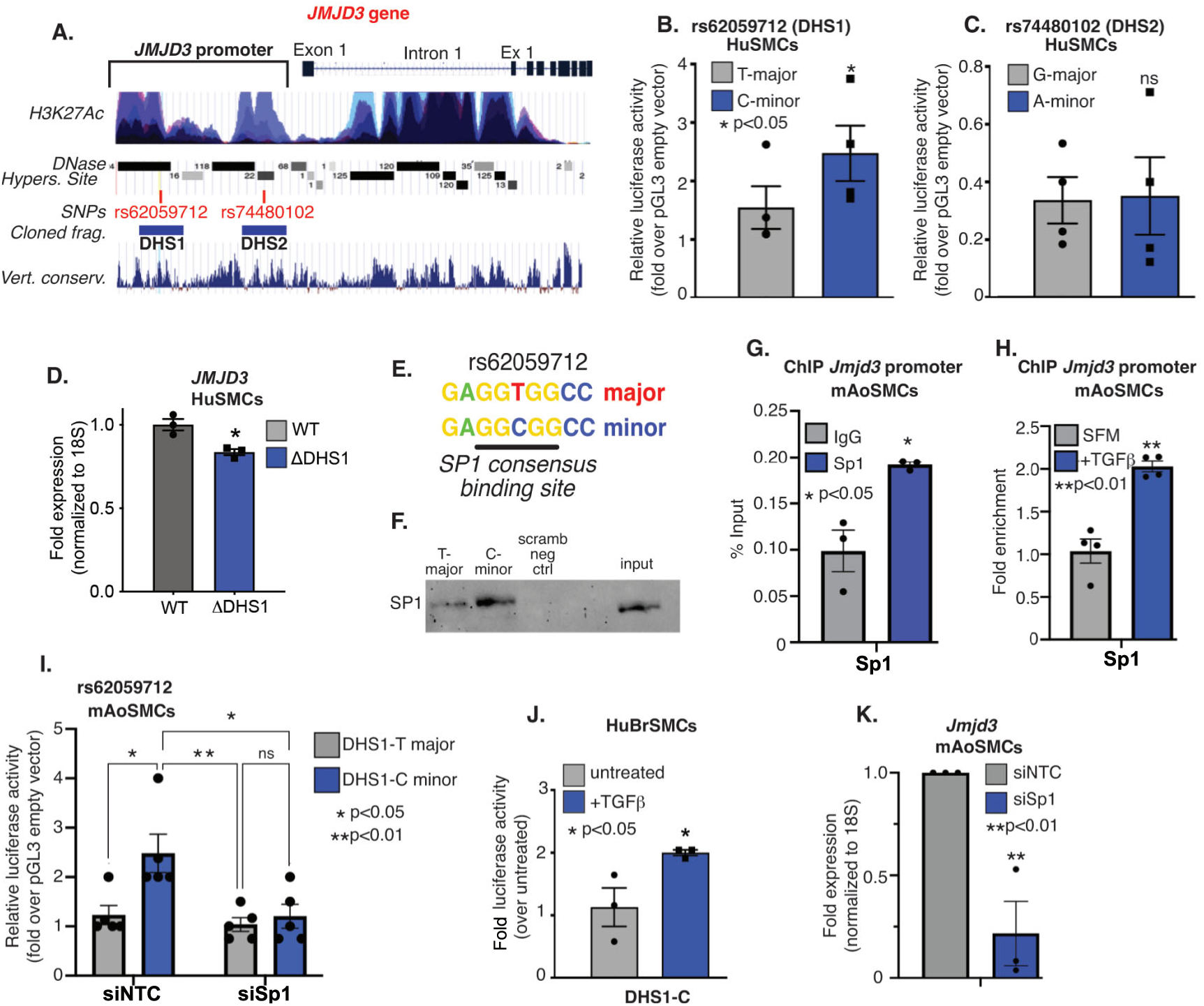
rs62059712 minor C allele increases JMJD3 transcription via enhanced SP1 binding. A) Schematic of the human *JMJD3* gene including gene structure, H3K27Ac, DHS, blood pressure-associated SNPs, cloned fragments for luciferase assays, and vertebrate sequence conservation obtained from UCSC genome browser. Features were used to prioritize SNPs in transcriptionally relevant regions. B) Luciferase results of the DHS1 containing rs62059712 (T-major vs. C-minor) and C) DHS2 containing rs74480102 (G-major vs. A-minor) in cultured primary human smooth muscle cells (HuSMCs). D) qPCR for *JMJD3* expression in HuSMCs with CRISPR-Cas9-mediated deletion of a 450 bp region encompassing the rs62059712 within DHS1 (ΔDHS1) compared with unedited (WT) HuSMCs. E) Sequence of the rs62059712 T-major and C-minor sequences with SP1 consensus binding site underlined. F) Western blot for SP1 after affinity purification using T vs. C probes corresponding to rs62059712 SNP region incubated with human aortic SMC (HuAoSMC) nuclear lysate. G) ChIP-qPCR for Sp1 at the *Jmjd3* promoter in mAoSMCs compared to IgG negative control. H) ChIP-qPCR for Sp1 at the *Jmjd3* promoter in mAoSMCs serum starved or treated with TGFβ. I) Luciferase assays in mAoSMCs treated with siNTC and siSp1 siRNAs then transfected with pGL3-DHS1-T and -C constructs. J) Luciferase assays in HuSMCs transfected with DHS1-C construct and then treated with TGFβ (20 ng/ml). K) qPCR for *Jmjd3* expression after NTC vs. Sp1 knockdown in mAoSMCs. Data are presented as the mean ± SEM. Results are representative of data from SMCs from 4-6 mice per group, n=3 independent experiments. Data were first analyzed for normal distribution, and if data passed the normality test, a two-tailed Student’s *t* test was used, *p<0.05, **p<0.01.

To define the upstream regulation whereby the rs62059712 minor C allele resulted in increased transcriptional activity, we analyzed the region of the *JMJD3* gene containing the rs62059712 C allele sequence for predicted transcription factor binding sites using the JASPAR web-based tool [39]. We found that the C containing sequence conformed to a predicted SP1 transcription factor binding site **(Figure 1E)**. Next, to examine whether SP1 bound the C allele in human SMCs, we performed affinity purification of transcription factors in HuSMC nuclear lysates using biotin-tagged DNA oligonucleotides that corresponded to the rs62059712 minor C, major T, or scrambled negative control sequences. In these experiments, the minor C allele bound the transcription factor, SP1 with greater affinity than the major T allele, supporting the predicted results from JASPAR **(Figure 1F)**. Because SP1 can interact with the transcription factor SMAD2 to drive TGFβ-dependent *ACTA2* expression in myofibroblasts, we also examined if the transcription factor SMAD2 could be immunoprecipitated with the C-minor allele probe [12]. As expected, we found increased SMAD2 binding to the C-containing sequence compared to the T-containing sequence **(Supplemental Figure 1C)**. Next, we examined by chromatin immunoprecipitation (ChIP) analysis in mAoSMCs if Sp1 was binding to the minor C allele and confirmed that Sp1 was present at the *Jmjd3* promoter **(Figure 1G)**. Furthermore, given that TGFβ is a well-known upstream driver of SMC differentiation, we assessed the effect of TGFβ treatment on Sp1 binding to the *Jmjd3* promoter [13, 40]. As demonstrated in **Figure 1H**, we observed increased Sp1 binding to the murine *Jmjd3* promoter in ChIP experiments following TGFβ stimulation of mAoSMCs. To determine if Sp1 was in fact mediating the increased transcription activity of the minor C allele, we transfected DHS1-T major and -C minor allele luciferase constructs in mAoSMCs that had been treated with siRNA against *Sp1* or a non-targeting control (NTC) siRNA. In support of our hypothesis that Sp1-mediated the allele-specific transcription activity of the DHS1 fragment, Sp1 knockdown in mAoSMCs abolished the increased transcriptional activity of the C minor allele, decreasing it to the activity of the T major allele **(Figure 1I)**. Additionally, since TGFβ increased SP1 binding to the *Jmjd3* promoter, we hypothesized that TGFβ may positively regulate transcription activity of the DHS1-C fragment. In support of this, TGFβ stimulation of mAoSMCs resulted in a 2-fold increase in luciferase activity over untreated DHS1-C fragment **(Figure 1J)**. Since the DHS1 region was required for *JMJD3* transcription (see Figure 1D) and SP1 binding to this region was important for its allele-specific activity in SMCs, we hypothesized that SP1 was important for *JMJD3* expression. Thus, to address this, we performed siRNA knockdown of Sp1 in mAoSMCs and then measured *Jmjd3* expression. Sp1 knockdown significantly decreased *Jmjd3* expression in mAoSMCs **(Figure 1K)**. Additionally, because SMAD2 also bound the *JMJD3* minor C allele, most likely via its interaction with SP1, we tested whether SMAD2 directly regulated *JMJD3* expression. siRNA knockdown of Smad2 also decreased *Jmjd3* expression in mAoSMCs **(Supplemental Figure 1D)**. Taken together, these results demonstrate that the blood pressure-associated human variant rs62059712 minor C allele increases *JMJD3* transcription in human and murine SMCs via increased SP1 binding.

### JMJD3 loss in vascular smooth muscle cells results in hypertension

Given our identification of a blood pressure-associated gene regulatory region within the human *JMJD3* promoter that displayed allele-specific activity in vascular SMCs, we hypothesized that JMJD3 was required for SMC-mediated vasomotor tone and HTN. To determine the role of JMJD3 in SMCs in blood pressure, we created a SMC-specific Jmjd3 deletion murine model by crossing our *Jmjd3^flox/flox^* mice (along with *Jmjd3^WT/WT^*and *Jmjd3^flox/WT^* mice) with tamoxifen-inducible *Myh11^CreERT^*mice (to generate *Jmjd3^flox/flox^Myh11^CreERT^* mice) **(Figure 2A)**. To eliminate the effect of tamoxifen on blood pressure and test the roles of heterozygous and homozygous *Jmjd3* deletion, we treated all *Jmjd3^WT/WT^Myh11^CreERT^*, *Jmjd3^flox/WT^Myh11^CreERT^*, and *Jmjd3^flox/flox^Myh11^CreERT^*littermates with tamoxifen for 5 days, followed by a 3-day “washout” period, to generate WT, *Jmjd3^flox/WT^Myh11^Cre+^*, and *Jmjd3^flox/flox^Myh11^Cre+^* mice, respectively. We then implanted osmotic minipumps (ALZET, Model 2004) filled with saline or Angiotensin II and measured blood pressure for 14 days in response to saline or Angiotensin II infusion (1 ug/kg/min). SMC-specific deletion of *Jmjd3* (in *Jmjd3^flox/flox^Myh11^Cre+^* mice) resulted in significantly higher systolic blood pressure (SBP), diastolic blood pressure (DBP), and mean arterial pressure (MAP) in response to Angiotensin II compared to heterozygote and wild-type controls **(Figures 2B-D)**. When blood pressures of Angiotensin II-treated mice were averaged over the course of the 14 day experiment, mice deficient in *Jmjd3* in SMCs had significant increases in SBP, DBP, and MAP compared to littermate controls **(Supplemental Figures 2A-C**).

**Figure 2.**
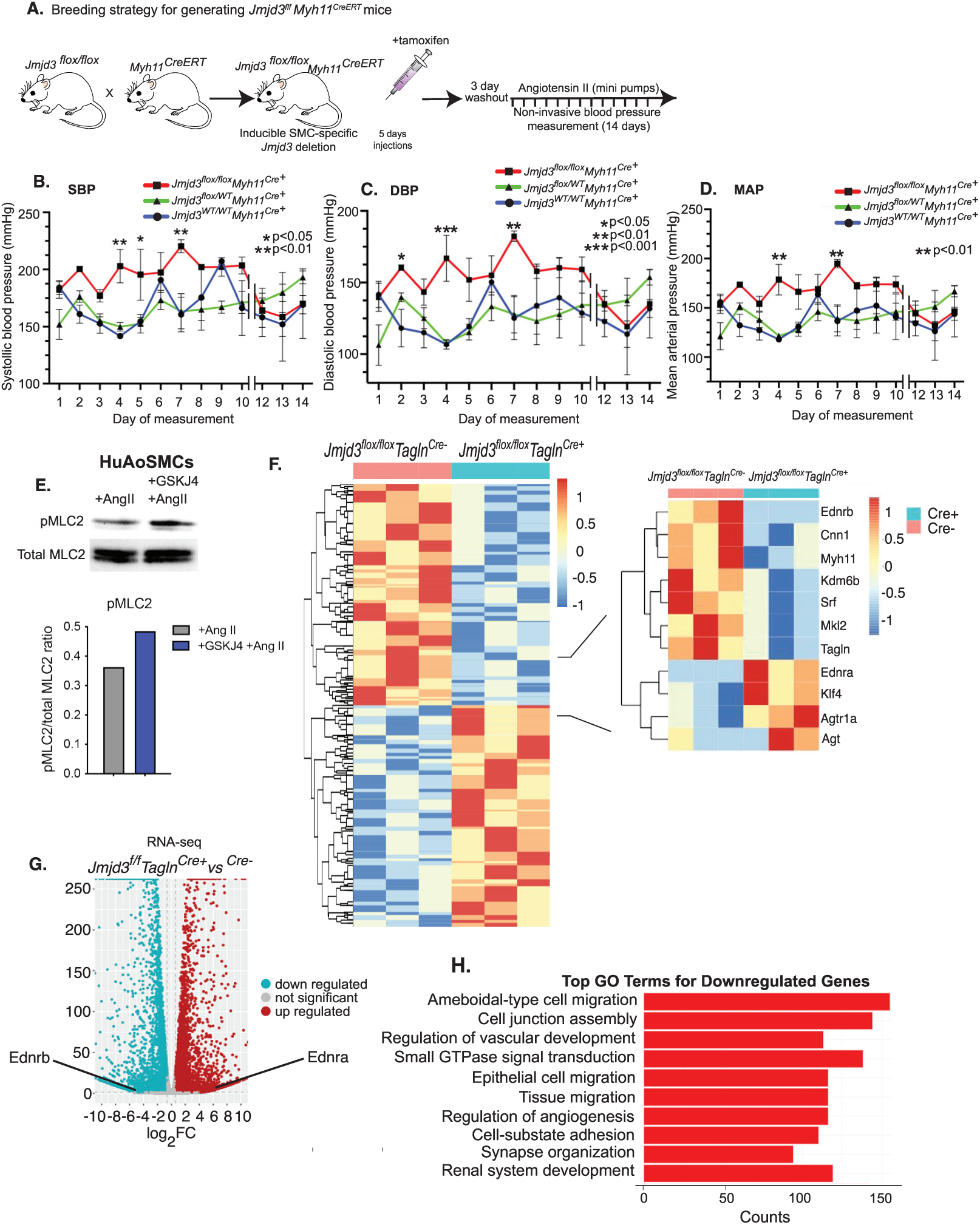
JMJD3 loss in vascular smooth muscle cells results in hypertension. A) Schematic depicting experiment and breeding strategy for generating inducible SMC-specific Jmjd3 knockout *Jmjd3^flox/flox^Myh11^CreERT^* mice. Blood pressure measurements obtained by tail cuff for 14 days in WT, heterozygous, and homozygous *Jmjd3^flox/flox^Myh11^CreERT^* mice treated with Angiotensin II infusion via osmotic minipumps. B) Systolic blood pressure (SBP), C) Diastolic blood pressure, (SBP) and D) Mean arterial pressure (MAP) are depicted. N=4-6 mice per genotype/group. E) HuAoSMCs were treated with Angiotensin II (100 nM) or Angiotensin II and GSKJ4 (50 nM) and then analyzed for phospho-myosin light chain 2 (pMLC2) and total MLC2 by western blotting. Blot is representative of n=3 independent experiments with representative densitometry depicted below. F) DEGs obtained from RNA-seq analysis of cultured mAoSMCs isolated from *Jmjd3^flox/flox^Tagln^Cre+^* and *Jmjd3^flox/flox^Tagln^Cre-^* mice with relevant DEGs depicted to right. DEGs depicted met significant threshold of p<0.05. Results obtained are representative of samples for each genotype submitted in triplicate and obtained from n=6-8 mice per sample. G) Volcano plot for upregulated (red) and downregulated (blue) DEGs with fold-change expression depicted on x-axis. Locations of *Ednrb* and *Ednra* are annotated. H) Bar graph of gene ontology (GO) analysis for top 10 downregulated genes in *Jmjd3^flox/flox^Tagln^Cre+^* SMC from RNA-seq results. Gene pathways listed on y-axis and number of gene counts for each pathway are listed on x-axis. Data are presented as the mean ± SEM, n=3 independent experiments for *in vitro* studies. Two-way ANOVA, *p<0.05, **p<0.01, ***p<0.001.

Since arterial SMCs are the main cell type regulating vascular tone and hence blood pressure, we hypothesized that JMDJ3 deletion resulted in increased vascular SMC contractility, thereby leading to increased arterial tone and blood pressure. Stimulation of SMC contraction occurs after activation of calcium/calmodulin-dependent phosphorylation of myosin light chain kinase, resulting in myosin light chain 2 (pMLC2) phosphorylation and smooth muscle contraction [3]. Thus, as a measure of SMC contractility, we measured pMLC2 via Western blotting and found it was increased in human aortic SMC (HuAoSMC) treated with a JMJD3 specific inhibitor (GSKJ4, 50 nM) plus Angiotensin II (100 nM) compared to Angiotensin II only (control) **(Figure 2E)**. To begin to elucidate the mechanism of JMJD3-dependent SMC contractility and blood pressure, we performed RNA sequencing (RNA-seq) on cultured *in vivo* aortic SMCs from *Jmjd3^flox/flox^Tagln^Cre+^* and *Jmjd3^flox/flox^Tagln^Cre-^*mice. As validation of Jmjd3 deletion in the *Jmjd3^flox/flox^Tagln^Cre^*model, *Jmjd3* mRNA levels in *Jmjd3^flox/flox^Tagln^Cre+^*SMCs were nearly 100% depleted **(Supplemental Figure 3A)**. Interestingly, among the many genes differentially regulated, we identified several canonical SMC-specific genes (*TAGLN, CNN1, MYH11, SRF, MKL2*) that were strongly downregulated in *Jmjd3^flox/flox^Tagln^Cre+^*SMCs **(Figure 2F)**. This was validated by decreased expression of SMC-specific marker proteins in aortic tissue from *Jmjd3^flox/flox^Tagln^Cre+^*mice **(Supplemental Figure 3B)**. Additionally, in this RNA-seq data, *Klf4*, a transcription factor that represses SMC differentiation, was upregulated in *Jmjd3^flox/flox^Tagln^Cre+^* SMCs compared to SMCs from *Jmjd3^flox/flox^Tagln^Cre-^* controls (see Fig. 2F). Given the important role of endothelin signaling in SMCs in blood pressure regulation, we examined endothelin receptor expression in our RNA-seq data and identified that there was a 6-fold downregulation in endothelin receptor B (*Ednrb*) expression and 6-fold upregulation in endothelin receptor A (*Ednra*) in *Jmjd3^flox/flox^Tagln^Cre+^* SMCs, suggesting this may be the underlying mechanism responsible for the HTN phenotype **(Figure 2G)**. As further evidence that JMJD3 in SMCs was important for vascular tone and blood pressure, other genes associated with HTN including *AGTR1A* and *AGT* were increased in *Jmjd3^flox/flox^Tagln^Cre+^* SMCs (see Fig. 2F). Notably, we performed gene ontology (GO) analysis using our RNA-seq results and identified common pathways related to migration, vascular development, angiogenesis, and signal transduction that were downregulated in *Jmjd3^flox/flox^Tagln^Cre+^* SMCs compared to control **(Figure 2H)**. Complementary GO analysis of upregulated genes in *Jmjd3^flox/flox^Tagln^Cre+^* SMCs revealed pathways common to DNA and RNA processes **(Supplemental Figure 3C)**. Taken together, these results demonstrate that deletion of JMJD3 in SMCs alters contractility, increases blood pressure, and leads to transcriptional changes in genes associated with SMC phenotype and hypertension.

### JMJD3 is required for EDNRB expression in SMCs and suppresses the hypertensive gene program

To further validate our RNA-seq results and perform a targeted analysis of downstream transcription targets of JMJD3 that drive blood pressure regulation in vascular SMCs, we performed a superarray of well-established genes involved in HTN in aortic SMCs isolated from *Jmjd3^flox/flox^Tagln^Cre^*mice. Notably, in further alignment with our RNA-seq data, there was nearly a 150-fold reduction in expression of the *Ednrb* gene in *Jmjd3^flox/flox^Tagln^Cre+^*SMCs as well as significant upregulation of multiple genes known to increase blood pressure (e.g., *Ednra, Agt, Ace2*) **(Supplemental Figure 4A)**. Results from the superarray were confirmed using targeted qPCR, which demonstrated decreased *Ednrb* expression (and increased expression of HTN-associated genes) in *Jmjd3^flox/flox^Tagln^Cre+^* SMCs **(Figure 3A, Supplemental Figures 4B-H**). To further examine the regulation of *EDNRB* expression by JMJD3, we performed siRNA knockdown of Jmjd3 in mAoSMCs and found a reduction in *Ednrb* expression compared to a non-targeting control (NTC) siRNA **(Figure 3B)**. In order to examine this *in vivo*, we harvested aortas from tamoxifen-injected *Jmjd3^flox/flox^Myh11^CreERT^*(*Jmjd3^flox/flox^Myh11^Cre+^*) and *Jmjd3^WT/WT^Myh11^CreERT^*(WT) mice after 14-day treatment with Angiotensin II (as described above in tail cuff blood pressure experiments) and found that *Ednrb* expression was significantly reduced in aortas from mice with SMC-specific *Jmjd3* deletion **(Figure 3C)**. Notably, there was an increase in Ednrb protein in aortic tissue in response to Angiotensin II that was abolished by SMC-specific *Jmjd3* deletion **(Figure 3D)**. The increase in *Ednrb* expression in aortas from mice treated with Angiotensin II compared to saline was also confirmed by qPCR **(Figure 3E)**. To determine whether the effects on *Ednrb* expression after Jmjd3 deletion were due to changes in H3K27me3, the epigenetic mark associated with Jmjd3, we performed ChIP for H3K27me3 at the *Ednrb* promoter. In order to examine this *in vivo*, we performed ChIP in mAoSMCs from *Jmjd3^flox/flox^Tagln^Cre+^*and *Jmjd3^flox/flox^Tagln^Cre-^* mice and observed increased H3K27me3 at the *Ednrb* promoter in *Jmjd3^flox/flox^Tagln^Cre+^* SMCs **(Figure 3F).** Additionally, we examined mAoSMCs treated with siRNA to *Jmjd3* or NTC siRNA and performed ChIP for H3K27me3 at the *Ednrb* promoter. In concordance with our above results, we found increased H3K27me3 at the *Ednrb* promoter following treatment with siRNA to *Jmjd3* compared to NTC siRNA **(Figure 3G)**. Next, to determine if the relationship between *JMJD3* and *EDNRB* expression was conserved in human SMCs, we isolated arteries from four HTN patients. scRNA-seq was performed and a Pearson correlation analysis identified that there was a significant correlation between the expression of these two genes in SMCs from human femoral arteries **(Figure 3H)**. Since there are two endothelin receptors (A and B) and prior reports have identified dual roles for these receptors, we examined if *Ednra* or *Ednrb* expression levels were altered relative to *Jmjd3* [3]. In both our superarray (see Supplemental Fig. 4A) and targeted qPCR in *Jmjd3^flox/flox^Tagln^Cre^* SMCs, we found increased *Ednra* expression in mAoSMCs with *Jmjd3* depletion **(Figure 3I)**. We also observed increased Ednra by immunofluorescent staining of *Jmjd3^flox/flox^Myh11^Cre+^*aortas compared to aortas from WT mice **(Figure 3J)**. Additionally, Ednra protein was increased in SMCs from *Jmjd3^flox/flox^Tagln^Cre+^* mice **(Figure 3K)**. Next, we performed siRNA-mediated knockdown of *Ednrb* in mAoSMCs, and then, to examine if *Ednrb* affected *Ednra* expression, measured *Ednra* expression by qPCR. Interestingly, Ednrb knockdown led to a 4-fold increase in *Ednra* expression in mAoSMCs, indicating that there is an intricate transcriptional balance between *EDNRA* and *EDNRB*, and hence, regulation of vessel contractility controlled by JMJD3 **(Figure 3L)**. Taken together, these results show JMJD3, likely via EDNRB, regulates SMC contractility and blood pressure.

**Figure 3.**
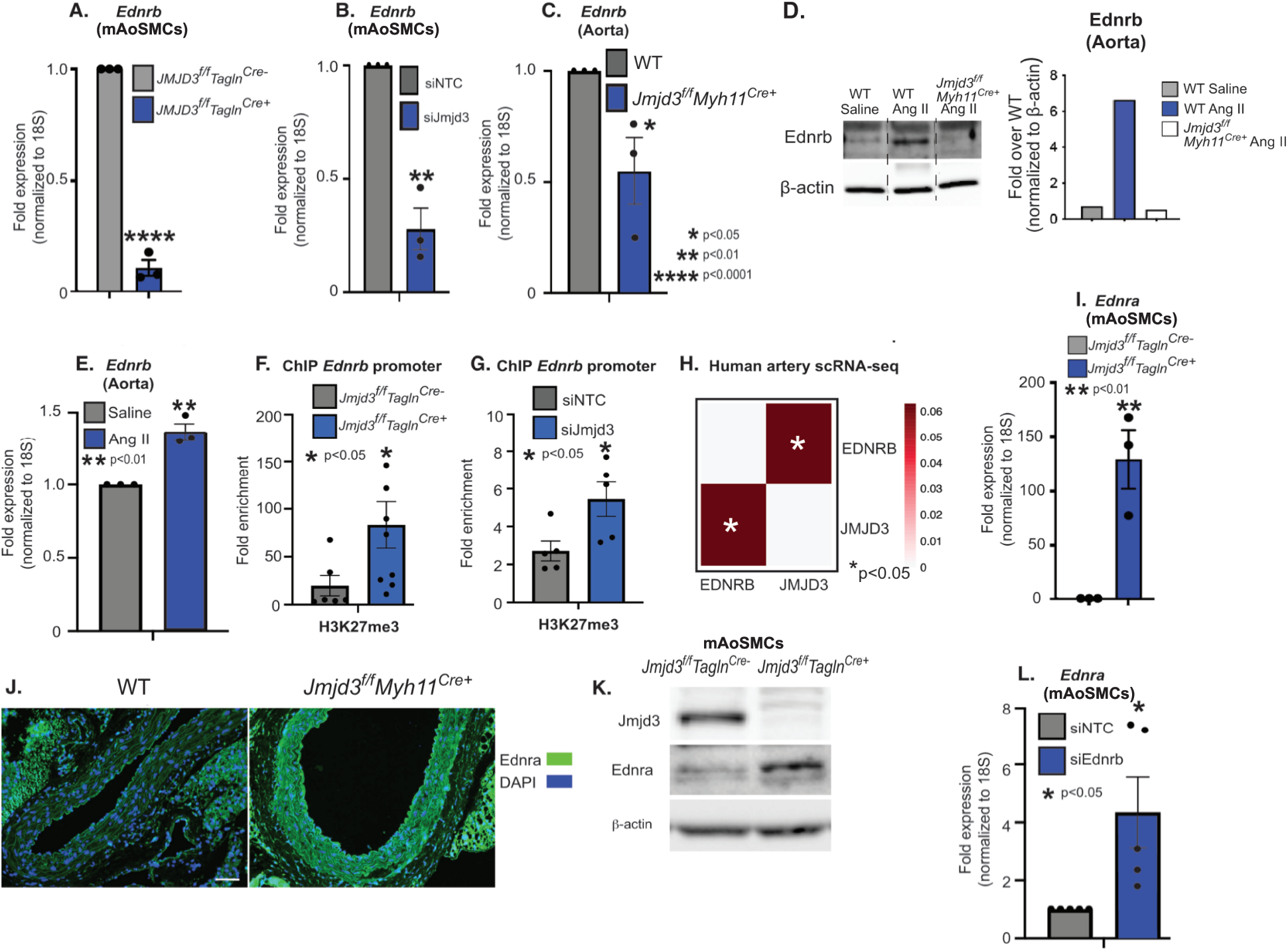
JMJD3 is required for *EDNRB* expression in SMCs and suppresses the hypertensive gene program. A) qPCR of *Ednrb* expression in SMCs isolated from *Jmjd3^flox/flox^Tagln^Cre+^* and *Jmjd3^flox/flox^Tagln^Cre-^* mice. B) qPCR for *Ednrb* in mAoSMCs treated with siNTC or siJmjd3 siRNA. C) qPCR of *Ednrb* expression in whole aortic tissue from *Jmjd3^flox/flox^Myh11^Cre+^* and WT aortas. D) Representative Western blot and densitometry results for Ednrb in aortas isolated from *Jmjd3^flox/flox^Myh11^Cre+^* and WT mice after 14 days of Angiotensin II or saline infusion. Representative densitometry is depicted to right. E) qPCR for *Ednrb* in aortas isolated from WT mice treated with saline or Angiotensin II for 14 days. F) ChIP-qPCR for H3K27me3 at the *Ednrb* promoter in mAoSMCs from *Jmjd3^flox/flox^Tagln^Cre^* mice. G) ChIP-qPCR for H3K27me3 at the *Ednrb* promoter in mAoSMCs treated with siNTC or siJmjd3 siRNA. H) scRNA-seq data from human femoral arteries showing Pearson correlation between *JMJD3* and *EDNRB* in SMCs, n=4 samples. I) qPCR of *Ednra* expression in SMCs isolated from *Jmjd3^flox/flox^Tagln^Cre^* mice. J) Representative images of immunofluorescent staining for Ednra in aortas harvested from *Jmjd3^flox/flox^Myh11^Cre+^* and WT mice after 14 days of angiotensin II infusion (1 ug/kg/min). Scale bar, 50 um. K) Representative western blotting results for Jmjd3 and Ednra in mAoSMCs from *Jmjd3^flox/flox^Tagln^Cre+^* or *Jmjd3^flox/flox^Tagln^Cre-^* mice. L) qPCR for *Ednra* in mAoSMCs treated with siNTC or siEdnrb siRNA. Data are presented as the mean ± SEM, n=3 independent experiments, n=4-6 mice per group. Data were first analyzed for normal distribution, and if data passed the normality test, a two-tailed Student’s *t* test was used, *p<0.05, **p<0.01, ***<0.0001.

### JMJD3 regulates vessel tone via endothelin-ERK signaling in vascular smooth muscle cells

Since blood pressure control is complex and regulated by multiple organ systems, we tested the effects of JMJD3 on vessel tone independent of organ tissues. To do this, we isolated aortic and mesenteric artery segments from *Jmjd3^flox/flox^Myh11^CreERT^* mice injected with tamoxifen (*Jmjd3^flox/flox^Myh11^Cre+^*) or corn oil (*Jmjd3^flox/flox^Myh11^Cre-^*) and measured their contractility using a vessel ring assay in response to various contractile agonists, including endothelin-1 (ET-1), angiotensin II (Ang II), and phenylephrine (PE), *ex vivo*. Since endothelin receptor signaling is a critical regulator of vascular SMC contractility and blood pressure, and *EDNRA* expression in SMCs was significantly upregulated after JMJD3 loss, we examined if increased *EDNRA* expression in vascular SMCs after JMJD3 loss resulted in increased vascular tone and HTN. Consistent with our above findings *in vivo* that SMC-specific JMJD3 loss resulted in HTN and that this was due to increased endothelin receptor expression, we observed that aortas isolated from *Jmjd3^flox/flox^Myh11^Cre+^* mice exhibited increased vessel tone compared to *Jmjd3^flox/flox^Myh11^Cre-^*littermate controls in response to ET-1 (10^-7^ M), and this was negated by treatment with the dual endothelin receptor antagonist, bosentan (10^-8^ M) **(Figure 4A)**. This was also observed in mesenteric arteries, and although *Jmjd3^flox/flox^Myh11^Cre+^*mesenteric arteries displayed higher baseline contractility than *Jmjd3^flox/flox^Myh11^Cre-^*littermate controls, there was no obvious difference in vessel contractility in response to Ang II or PE **(Figure 4B)**. This indicated that the effects of JMJD3 deletion on increased vessel tone were specific to endothelin signaling rather than other vasoactive agonists. In agreement with this, *Jmjd3^flox/flox^Myh11^Cre+^* aortas exhibited increased responsiveness to ET-1 stimulation compared to aortas isolated from *Jmjd3^flox/flox^Myh11^Cre-^*controls, and this enhancement in contractility was reduced to *Jmjd3^flox/flox^Myh11^Cre-^* baseline levels following bosentan treatment **(Figure 4C)**.

**Figure 4.**
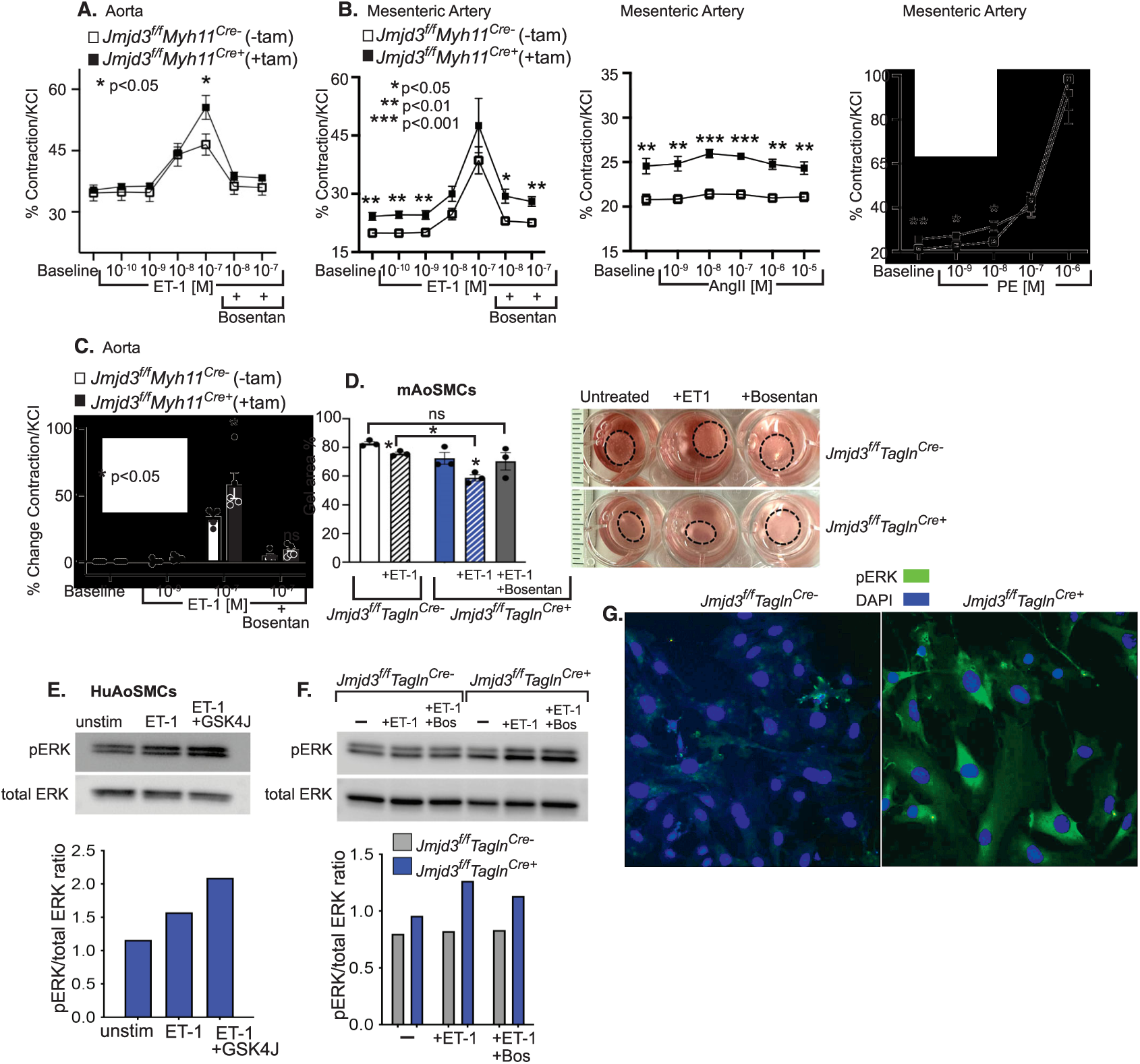
JMJD3 regulates vessel tone via endothelin-ERK signaling in vascular smooth muscle cells. A) *Jmjd3^flox/flox^Myh11^CreERT^* mice were injected with tamoxifen (*Jmjd3^flox/flox^Myh11^Cre+^*) or corn oil (*Jmjd3^flox/flox^Myh11^Cre-^*) and then aortas were harvested, suspended on wire hooks, basal tone normalized with KCl, and then treated with increasing concentrations of endothelin-1 (ET-1, 0-10^-7^ M). Once maximal response was observed, vessel segments were treated with bosentan (10^-8-^10^-7^ M) plus ET-1 at dose (10^-7^ M) that elicited maximal contraction. B) Mesenteric arteries were isolated as in A) and suspended on wires and normalized vessel tone was obtained. Mesenteric artery segments were treated with increasing concentrations of ET-1 (0-10^-7^ M) and bosentan (10^-8-^10^-7^ M) plus dose of ET-1 (10^-7^ M) that elicited maximal contractile response, Angiotensin II (Ang II, 0-10^-5^ M), or Phenylephrine (PE, 0-10^-6^ M). C) Graph depicting contractility of aortic rings treated with ET-1 (0-10^-7^ M) plus or minus bosentan (10^-7^ M) as percent change in contraction relative to baseline contractility. D) Cultured mAoSMCs from *Jmjd3^flox/flox^Tagln^Cre^* mice were embedded in collagen gels that were allowed to polymerize and either untreated or treated with ET-1 (1 uM), ET-1 plus bosentan (10 uM), or bosentan alone. Results are depicted as percentage of initial gel area at 24 hours after seeding in collagen gels. E) Representative western blot of HuAoSMCs unstimulated, treated with ET-1 (1 uM), or ET-1 plus GSKJ4 (50 nM) and then probed for phosphorylated ERK (pERK, top) or total ERK (bottom). Blot with representative densitometry results underneath. F) Representative western blot of *Jmjd3^flox/flox^Tagln^Cre+^* and *Jmjd3^flox/flox^Tagln^Cre-^* control mAoSMCs unstimulated, treated with ET-1 (1 uM), or ET-1 plus bosentan (10 um) and then probed for pERK (top) or total ERK (bottom). Blot with representative densitometry results underneath. G) Representative immunofluorescent staining for pERK in *Jmjd3^flox/flox^Tagln^Cre+^* (right) and *Jmjd3^flox/flox^Tagln^Cre-^* (left) mAoSMCs (up to 4 high powered fields counted per experiment). Data are presented as the mean ± SEM, n=3 independent experiments for *in vitro* studies, n=6 mice per group for contractility experiments. *In vitro* experiments representative of SMCs from 4-6 mice per group. Two-way ANOVA and two-tailed Student’s t-test were used. *p<0.05, **p<0.01, ***p<0.001.

To interrogate the role of SMCs specifically in the altered vessel contractile response and as another model for testing SMC contractility, we utilized a collagen gel contraction assay in which SMCs from *Jmjd3^flox/flox^Tagln^Cre^* mice were isolated *in vivo* and then embedded in a collagen gel, treated with ET-1 (1 uM) and/or bosentan (10 uM), and gel area was measured. We identified that gels containing *Jmjd3* deficient mAoSMCs had smaller gel areas (associated with increased SMC contraction) compared to *Jmjd3^flox/flox^Tagln^Cre-^* SMCs in response to ET-1, and treatment of gels containing SMCs deficient in *Jmjd3* with bosentan resulted in gel areas comparable to untreated gels **(Figure 4D)**. Taken together, these results suggest that increased endothelin signaling associated with JMJD3 deletion results in increased SMC contraction, leading to higher arterial tone and elevations in blood pressure.

Endothelin receptor stimulation leads to ERK pathway activation, which has an established role in increasing SMC contraction [3, 41, 42]. In concordance with this, we found that treatment of HuAoSMCs with ET-1 (1 uM) for 5 minutes led to an increase in pERK by Western blotting **(Figure 4E)**. Pre-treatment of HuAoSMC with the JMJD3 inhibitor GSKJ4 (50 nM) led to an increased pERK signal in response to ET-1, suggesting that enhanced endothelin signaling after JMJD3 loss in SMCs results in increased ERK signaling. To further interrogate this, we treated mAoSMCs from *Jmjd3^flox/flox^Tagln^Cre^*mice with ET-1 (1 uM) with or without the endothelin receptor antagonist bosentan (10 uM) and then measured pERK by Western blotting. As shown in **Figure 4F**, baseline ERK activity was modestly increased in Jmjd3-deficient SMCs compared to *Jmjd3^flox/flox^Tagln^Cre-^* control SMCs, and treatment with ET-1 resulted in a substantial increase in pERK in Jmjd3-deficient SMCs compared to *Jmjd3^flox/flox^Tagln^Cre-^*controls. Pre-treatment of mAoSMCs with bosentan resulted in a small reduction in pERK activity in Jmjd3-deficient mAoSMCs. In order to further examine ERK activity, we used immunofluorescence and observed increased pERK in *Jmjd3^flox/flox^Tagln^Cre+^* SMCs absent in Jmjd3 compared to *Jmjd3^flox/flox^Tagln^Cre-^* control SMCs **(Figure 4G)**. Taken together, these results demonstrate that JMJD3 regulates endothelin-pERK signaling in SMCs.

### JMJD3 is required for vascular smooth muscle cell differentiation

One hallmark associated with long-standing HTN is the transition of SMCs from a contractile to a synthetic phenotype [43, 44]. Given our findings that JMJD3 in SMCs altered blood pressure *in vivo*, we investigated if SMC phenotype is altered by JMJD3. We examined mAoSMCs for *Jmjd3* expression after stimulation with TGFβ (20 ng/ml), since TGFβ is a well-established driver of SMC differentiation [13]. We found that TGFβ increased *Jmjd3* expression in mAoSMCs by approximately 2-fold **(Figure 5A)**. Next, we performed ChIP for Jmjd3 on mAoSMCs at canonical SMC-specific gene promoters (*Acta2, Tagln, Cnn1, Myh11*) that control SMC plasticity. Jmjd3 demonstrated significant enrichment at smooth muscle gene promoters in mAoSMCs **(Figure 5B)**. We then used siRNA-mediated knockdown of Jmjd3 in mAoSMCs treated with TGFβ (20 ng/ml) to investigate the effect of Jmjd3 on SMC-specific gene expression (*Acta2, Tagln, Cnn1, Myh11*). Jmjd3 knockdown with siRNA reduced TGFβ-dependent expression of SMC genes (*Acta2, Tagln, Myh11*) at the mRNA and protein levels compared to control NTC siRNA **(Figures 5C, D)**. To examine this genetically, smooth muscle gene expression was significantly reduced in *Jmjd3^flox/flox^Tagln^Cre+^* SMCs compared to *Jmjd3^flox/flox^Tagln^Cre-^* control SMCs **(Figure 5E)**. In order to test this pharmacologically, treatment of mAoSMCs with the Jmjd3 selective inhibitor GSKJ4 (50 nM) inhibited smooth muscle gene expression (*Acta2, Tagln, Cnn1, Myh11*) **(Figure 5F)**. In order to examine the direct effects of Jmjd3 on SMC gene promoters, ChIP was performed for H3K27me3 on mAoSMCs treated with siRNA to *Jmjd3* or NTC siRNA. Jmjd3 knockdown with siRNA resulted in increased H3K27me3 enrichment at SMC-specific gene promoters (*Acta2, Tagln, Cnn1, Myh11*), indicating that JMJD3 positively regulates SMC gene expression directly by removing H3K27me3 from SMC gene promoters **(Figure 5G-J)**. These results reveal that *JMJD3* expression is increased by TGFβ and is required for SMC gene expression via an H3K27me3-mediated mechanism.

**Figure 5.**
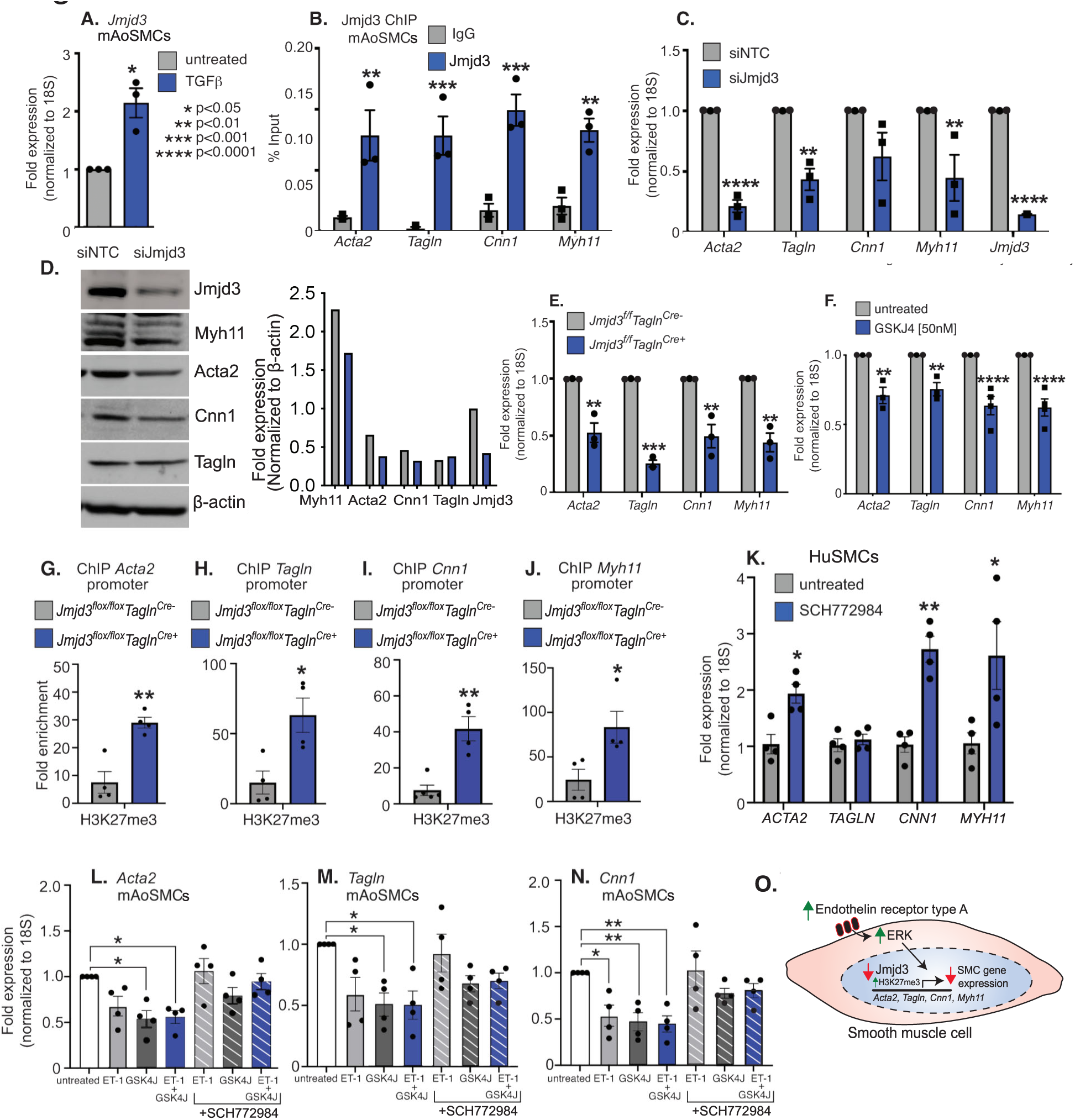
JMJD3 is required for vascular smooth muscle cell differentiation. A) *Jmjd3* expression in mAoSMCs untreated versus treatment with TGFβ (20 ng/ml). B) ChIP-qPCR showing Jmjd3 enrichment at SMC gene promoters compared to a negative control IgG. C) SMC gene expression in mAoSMCs treated with Jmjd3 siRNA or NTC siRNA for 72 hours total, serum starved, and then treated with TGFβ (20 ng/ml). D) Representative western blot probed for SMC markers in lysates from mAoSMCs treated with Jmjd3 siRNA or NTC siRNA. Representative densitometry of blot is depicted to right. E) Smooth muscle gene expression in SMCs cultured from *Jmjd3^flox/flox^Tagln^Cre+^* mice and *Jmjd3^flox/flox^Tagln^Cre-^* littermate controls. F) Expression of *Acta2, Tagln, Cnn1, and Myh11* in mAoSMCs treated with Jmjd3 inhibitor GSKJ4 (50 nM) for 16 hours. G) H3K27me3 ChIP-qPCR at *Acta2*, *Tagln* (H), *Cnn1* (I), *Myh11* (J) promoters in *Jmjd3^flox/flox^Tagln^Cre^* mAoSMCs. K) qRT-PCR for SMC markers in HuSMCs treated with SCH772984 (5 uM) for 16 hours compared to untreated. L) qRT-PCR for *Acta2*, *Tagln* (M), and *Cnn1* (N) in mAoSMCs treated with ET-1 (1 uM), GSK4J (50 nM), ET-1 with GSK4J with or without the ERK inhibitor, SCH772984 (5 uM). O) Illustration depicting the effect of JMJD3 loss on SMC gene expression due to both increased EDNRA and ERK activation as well as increased H3K27me3 at SMC gene promoters. Data are presented as the mean ± SEM, n=3 independent experiments. Tissues harvested from n=4-6 mice per group. *In vitro* experiments representative of SMCs from 4-6 mice per group. Two-way ANOVA and two-tailed Student’s t-test were used. *p<0.05, **p<0.01, ***p<0.001, ****p<0.0001.

Given our findings that JMJD3 loss in SMCs resulted in increased expression of *ENDRA*, the predominant endothelin receptor in SMCs, leading to increased endothelin signaling and ERK activation and evidence from others demonstrating that increased ERK activity results in repression of SMC gene expression and phenotypic modulation, we explored whether JMJD3 controlled SMC gene expression by regulating ERK signaling [45]. First, as a translational corollary, treatment of human SMCs with the ERK inhibitor SCH772984 (5 uM) robustly increased SMC gene expression (*ACTA2, CNN1, MYH11*) **(Figure 5K)**. Next, to examine the effect of ERK inhibition in the setting of JMJD3 loss, we treated mAoSMCs with combinations of GSKJ4 (50 nM), ET-1 (1 uM), and/or SCH772984 (5 uM). Consistent with our hypothesis, ET-1 treatment led to a significant reduction in SMC gene expression, which was further reduced by inhibiting Jmjd3 with GSKJ4 (50 nM). Supporting the role of ERK in repressing SMC gene expression after Jmjd3 inhibition, treatment of mAoSMCs with SCH772984 (5 uM) prevented downregulation of SMC gene expression by ET-1 (1 uM) and GSKJ4 (50 nM) alone and in combination with each other **(Figure 5L-N)**. These data reveal a dual-mechanism for JMJD3 on SMC gene transcription whereby JMJD3 loss results in decreased smooth muscle gene expression via increased H3K27me3 at SMC gene promoters and by increased endothelin-ERK activation, inhibition of which restores SMC gene expression **(Figure 5O)**.

### Hypertensive-induced arterial remodeling is regulated by JMJD3

Since our data indicate that JMJD3 is a regulator of SMC differentiation and the endothelin/ERK signaling pathway, which independently controls SMC phenotype, we examined the role of JMJD3 in SMCs on arterial remodeling seen during long-standing hypertension. Given that blood pressure is regulated by resistance arteries, we measured the renal arteriole wall thickness from *Jmjd3^flox/flox^Myh11^Cre+^* and WT mice treated with Angiotensin II for 14 days and observed increased media to diameter ratio in mice with SMC-specific deletion of Jmjd3 compared to littermate controls, indicating increased remodeling in the absence of Jmjd3 in SMCs **(Figure 6A, B)**. Since Angiotensin II induces SMC phenotypic modulation and arterial remodeling, we explored whether Angiotensin II regulates JMJD3 expression, thereby leading to changes in SMC gene expression during HTN [43, 46]. We first measured *Jmjd3* mRNA in aortas isolated from mice treated with saline control or Ang II and observed decreased *Jmjd3* expression in the aortas isolated from Ang II-treated mice **(Figure 6C)**. This was accompanied by a reduction in expression of *Acta2, Tagln, Cnn1, and Myh11* measured in whole aorta tissue **(Figures 6D-G)**. Given that JMJD3 was required for SMC differentiation (see Figure 5), we examined if JMJD3 also regulated SMC gene expression during Angiotensin II-mediated hypertensive arterial remodeling. We measured smooth muscle gene expression (*Acta2, Tagln, Cnn1, Myh11*) in *Jmjd3^flox/flox^Myh11^Cre+^* and WT mice that were treated with Angiotensin II for 14 days. SMC-specific Jmjd3 deletion resulted in further loss of SMC markers in aortas from mice treated with Angiotensin II at the protein and mRNA levels **(Figures 6H-L)**. Since KLF4 is a critical transcription factor that controls the switch of SMCs from the mature, contractile state to the proliferative, synthetic phenotype, we examined if *KLF4* expression was altered under hypertensive conditions [14, 16]. Indeed, Angiotensin II (100 nM) increased *KLF4* expression in HuAoSMCs, suggesting that KLF4 may drive phenotypic modulation during hypertensive remodeling **(Figure 6M)**. Because loss of JMJD3 promoted the synthetic SMC phenotype, we tested if KLF4 was transcriptionally regulated by JMJD3. First, we assessed *Klf4* expression in SMCs from *Jmjd3^flox/flox^Tagln^Cre+^* and *Jmjd3^flox/flox^Tagln^Cre-^* littermate controls and demonstrated a significant 2.5-fold upregulation in *Klf4* expression in the setting of Jmjd3 deletion in *Jmjd3^flox/flox^Tagln^Cre+^* cultured SMCs **(Figure 6N)**. Additionally, siRNA knockdown of Jmjd3 in mAoSMCs increased *Klf4* expression nearly 3-fold compared to NTC siRNA **(Figure 6O)**. Since SMC migration is regulated by ERK signaling and phenotypic modulation, which are both regulated by Jmjd3, we assessed migration using scratch assays with *Jmjd3^flox/flox^Tagln^Cre+^*and *Jmjd3^flox/flox^Tagln^Cre-^* SMCs. Jmjd3-deficient *Jmjd3^flox/flox^Tagln^Cre+^* SMCs exhibited increased migration speed compared to *Jmjd3^flox/flox^Tagln^Cre-^*control SMC **(Figure 6P)**. In summary, Angiotensin II-induced downregulation of *JMJD3* in SMCs results in loss of mature SMC genes and increased *KLF4* expression, which cooperatively drive phenotypic modulation and remodeling during long-standing HTN **(Figure 6Q).**

**Figure 6.**
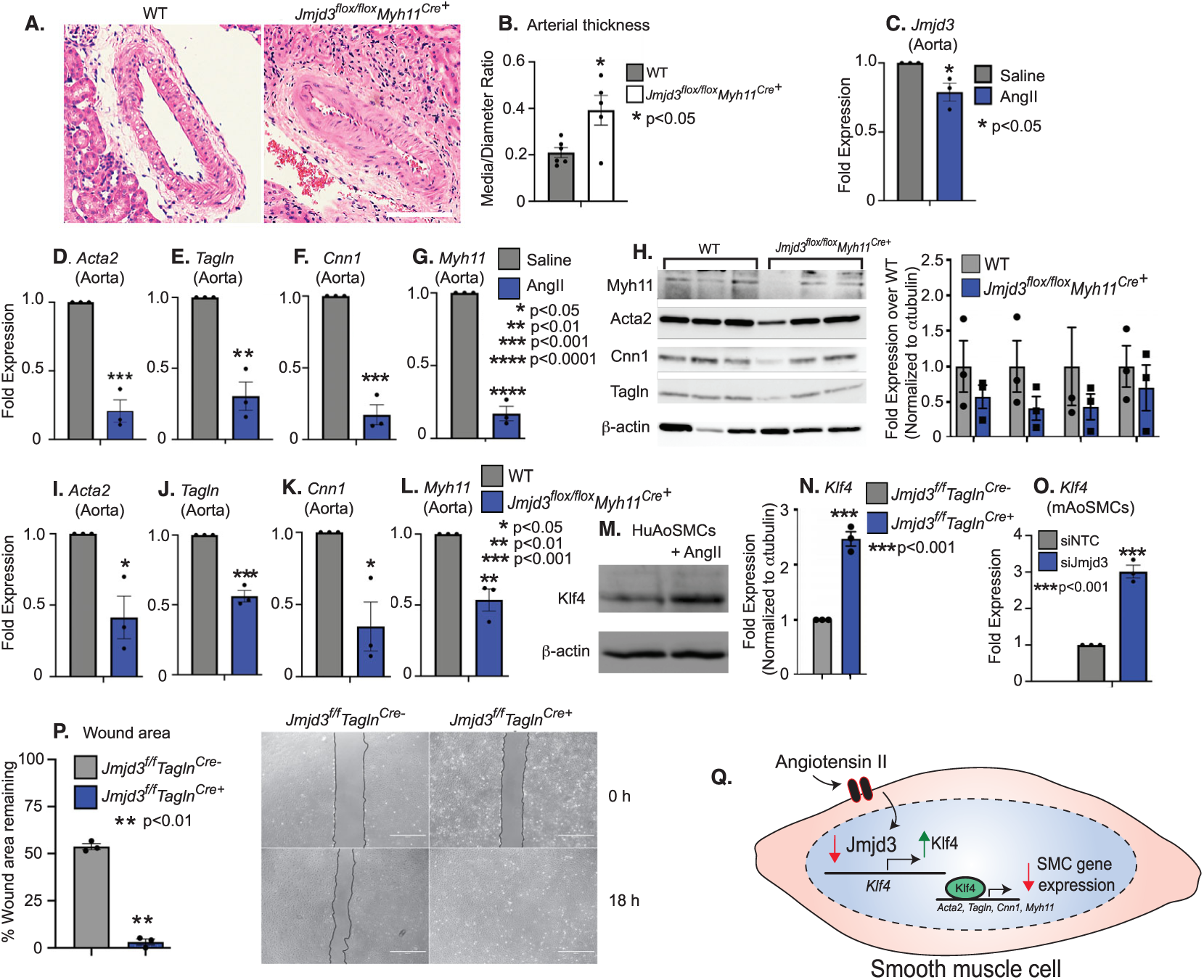
Hypertensive-induced arterial remodeling is regulated by JMJD3. A) H&E staining of kidney sections showing renal arterioles from *Jmjd3^flox/flox^Myh11^Cre+^* and WT mice treated with Angiotensin II infusion (1 ug/kg/min) for 14 days. Scale bar, 100 um. B) Measurement of renal arteriole medial wall thickness from (A). C) qPCR for *Jmjd3* in WT aortas from mice treated with 14 days of Angiotensin II or saline infusion. D) qPCR for *Acta2, Tagln* (E), *Cnn1* (F), *Myh11* (G) in aortas from WT mice treated with 14 days of Angiotensin II or saline infusion. H) Western blot for SMC markers from whole aortas from *Jmjd3^flox/flox^Myh11^Cre+^* and WT mice with densitometry to right. I) qPCR for *Acta2, Tagln* (J), *Cnn1* (K), and *Myh11* (L) in aortas from *Jmjd3^flox/flox^Myh11^Cre+^* and WT mice treated with 14 days of Angiotensin II. M) Representative western blot probed for KLF4 in HuAoSMCs serum starved or treated with Angiotensin II for 16 hours. Blot representative of n=3 independent experiments. N) qPCR analysis of *Klf4* expression in *Jmjd3^flox/flox^Tagln^Cre^* mAoSMCs. O) qPCR analysis of *Klf4* expression in mAoSMCs transfected with NTC or siJmjd3 siRNAs. P) Results of scratch assay on *Jmjd3^flox/flox^Tagln^Cre^* mAoSMCs as percent wound area remaining after 18 hours following initial scratch. Representative images at 0 hours and 18 hours after scratch are depicted on right. Scale bar, 1 mm. Q) Schematic of transcriptional regulation of Angiotensin II-JMJD3-KLF4 axis controlling SMC gene expression. Data are presented as the mean ± SEM, n=3 independent experiments. Tissues harvested from n=4-6 mice per group. *In vitro* experiments representative of SMCs from 4-6 mice per group. Two-tailed Student’s t-test was used. *p<0.05, **p<0.01, ***p<0.001, ****p<0.0001.

### JMJD3 regulates the contractile gene program in SMCs by cooperatively regulating SRF binding to SMC gene promoters

We showed above that JMJD3 regulates smooth muscle response to Angiotensin II-induced HTN, and loss of JMJD3 results in decreased SMC gene expression and increased pathologic arterial remodeling. In order to translate these *in vivo* murine findings to humans, we performed single cell RNA-sequencing (scRNA-seq) in human femoral artery samples (n=4) **(Figure 7A)**. First, we measured *JMJD3* expression in SMCs from human arteries and found that *JMJD3* was expressed in SMCs at moderate levels **(Figure 7B)**. Next, we performed Pearson expression correlation analysis between genes associated with contractile and synthetic gene programs in SMCs. As shown in **Figure 7C**, genes associated with mature, contractile SMC phenotype including *ACTA2, TAGLN, CNN1,* and *MYH11* showed very strong correlation with one another, yet weak or no correlation with synthetic, proliferative-associated genes including *PDGFBR, PDGFB, KLF4,* and *ETS-1*. Similarly, the proliferative genes exhibited strong correlation with each other. The distinct clustering between the synthetic and contractile gene sets confirmed the utility of this approach. Next, to determine how JMJD3 transcriptionally interacts with the SMC contractile gene program, we performed analysis of relative *JMJD3* expression in SMC subsets separated into high or low expression of contractile genes. We found that *JMJD3* expression (as was the number of JMJD3-expressing cells) was increased in *ACTA2* and *CNN1* “high” SMCs compared to “low”-expressing SMCs **(Figure 7D)**, further supporting the role of JMJD3 as a positive regulator of the contractile smooth muscle gene program. Because chromatin modifying enzymes cooperatively regulate transcription factor binding, we investigated if JMJD3 interacted with serum response factor (SRF), a transcription factor essential for driving SMC gene expression. Using proximity ligation assay, we demonstrated physical interaction between Jmjd3 and Srf in the nuclei of mAoSMCs **(Figure 7E)**. We also performed co-immunoprecipitation in mAoSMC nuclear lysates and observed a weak interaction between Jmjd3 and Srf **(Supplemental Figure 5)**. Finally, to determine the functional consequence of Jmjd3 on Srf binding to DNA, we performed ChIP for Srf at SMC gene promoters on mAoSMCs treated with *Jmjd3* siRNA or NTC siRNA. We observed decreased binding of Srf to *Acta2*, *Tagln*, and *Cnn1* promoters in Jmjd3 knockdown mAoSMCs treated with *Jmjd3* siRNA compared to NTC siRNA **(Figure 7F)**. Taken together, our data reveal that JMJD3 regulates the contractile gene program in human SMCs and establish a role for JMJD3 in cooperatively regulating SRF binding at smooth muscle gene promoters.

**Figure 7.**
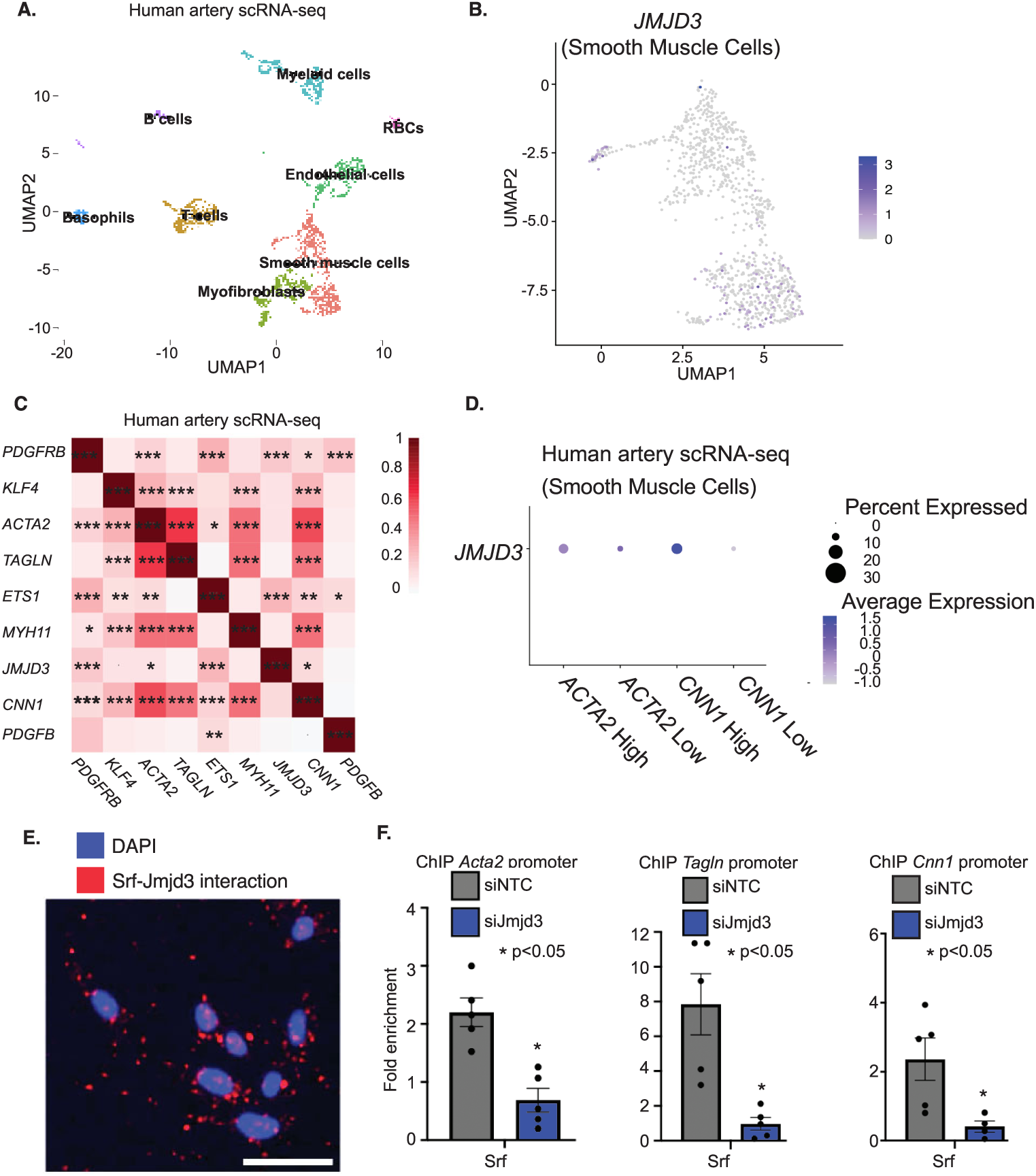
JMJD3 regulates the contractile gene program in SMCs by cooperatively regulating SRF binding to SMC gene promoters. A) UMAP plots showing distribution of cell populations from human scRNA-seq of human femoral artery samples (n=4 samples). B) Relative expression of *JMJD3* in SMCs in human artery. C) Graphical representation of Pearson correlation of smooth muscle genes obtained from single cell RNA-seq of human arteries with heat gradient representing strength of association. D) Dot plot of *JMJD3* expression in human artery SMCs separated by high versus low expression of *ACTA2* and *CNN1*. E) Representative image of PLA assay in mAoSMCs demonstrating interaction between Srf and Jmjd3 (red foci). At least 4 high powered fields were analyzed. Scale bar, 50 um. F) Srf ChIP-qPCR using primer sets for the *Acta2*, *Tagln*, and *Cnn1* promoters in mAoSMCs treated with NTC or siJmjd3 siRNA. Data are presented as the mean ± SEM, n=3 independent experiments. N=4 arterial samples. Experiments representative of SMCs from 4-6 mice per group. Two-tailed Student’s t-test and Pearson’s correlation coefficient were used. *p<0.05, **p<0.01, ***p<0.001.

## Discussion

Here, we define a causal role for the blood pressure-associated polymorphism rs62059712 in the transcriptional regulation of *JMJD3*, which we show regulates endothelin-dependent blood pressure and smooth muscle cell phenotype via epigenetic alterations at SMC gene promoters as well as via changes in ERK signaling. This work uncovers an allele-specific mechanism regulating expression of an epigenetic enzyme (JMJD3) that controls expression of the endothelin receptors, and consequently, modulates SMC contractility, differentiation, and arterial remodeling in response to long-standing hypertension. As such, these findings identify a novel pathway linking genetic and epigenetic mechanisms to molecular control of SMC function and phenotype, and in doing so, reveal important mechanistic insights into how GWAS and similar studies can be used to identify and define novel epigenetic regulators of disease. Our findings indicate the potential of using rs62059712 genotype for identifying therapeutic response to specific anti-hypertensive medications, which may also alleviate the pathologic consequences of hypertension (e.g., arterial remodeling) in select patients.

We characterize the effect of the blood pressure-associated SNP rs62059712 on *JMJD3* transcription, demonstrating that the minor C allele increases the activity of this regulatory region by creating an SP1 binding site within the *JMJD3* promoter. SP1 plays important roles in regulating SMC differentiation and phenotype [12, 14]. SP1 is generally reported as a repressive transcription factor that induces the SMC phenotypic switch after vascular injury by directly binding to GC repressor elements in SMC-specific genes (e.g., *MYH11*) as well as indirectly by increasing expression of *KLF4*, which inhibits myocardin function and downstream SMC differentiation. Notably, in these studies, SP1 was activated by upstream signals (e.g., PDGF) and in disease contexts that induce SMC phenotypic modulation, however, regulation of basal SMC gene transcription by SP1 was not explored. In contrast, reports have also demonstrated positive effects of SP1 on gene transcription, for example, increasing *ACTA2* expression in myofibroblasts [12]. Furthermore, interactions involving other epigenetic complexes, such as p300 and acetylated histone 3, cooperatively regulate SP1 function and downstream SMC function [47]. We show that the DHS1 C-minor allele also interacts with SMAD2, which is required for *JMJD3* expression in SMCs. As such, it is likely that SP1 in combination with SMAD2 and other yet-to-be identified transcription factors positively regulates *JMJD3* transcription. Whether or not SMAD2 physically interacts with SP1 and/or directly binds to the C-minor allele DNA sequence was not explored in this study. Additionally, binding partners of JMJD3 that regulate its activity and SMC-specific gene expression need to be explored. We have begun to analyze some of these potential interactions, demonstrating that JMJD3 interacts with SRF and is required for its binding at SMC gene promoters. Identifying additional JMJD3 binding partners will provide further insight into epigenetic regulation of SMC phenotype in disease.

Blood pressure regulation is controlled by numerous upstream molecular, genetic, and epigenetic signals. The utility of GWAS and other large-scale studies in identifying true gene-disease associations is obscured by the saturation of non-causal mutations in these studies. Despite this, functional mutations affecting blood pressure have been identified by GWAS that have provided insight into the genetic regulation of blood pressure [48, 49]. Of particular interest to the current study, at least three SNPs have been identified that influence endothelin signaling, indicating the genetic complexity of endothelin pathway. rs9349379 in the intron of the *PHACTR1* gene affects *ET1* expression by altering long-range interactions between the *PHACTR1* and *ET1* gene loci [50]. Two other SNPs, rs1630736 in the *ET1* gene, and rs10305838 in the *EDNRA gene*, are also associated with blood pressure [51, 52]. Here, we find that JMJD3 loss in SMC leads to increased endothelin-ERK signaling, resulting in decreased SMC gene expression. By demonstrating that ERK inhibition reverses this effect, we corroborate previous studies that implicate the ERK pathway in regulating smooth muscle phenotype [45]. Additionally, our results suggest that targeting ERK in long-standing hypertension, and perhaps other models of vascular injury, may limit the development of pathologic arterial remodeling. Giri et al. reported a 0.39 mmHg increase systolic blood pressure for each copy of the major allele [30]. While this per copy allele effect is seemingly small, albeit typical for similar reported SNPs, it is likely that this SNP interacts with other genetic, epigenetic, and molecular cues that all function in a mosaic fashion to create larger changes in blood pressure.

While prior studies have investigated the role of epigenetic alterations in cardiovascular disease, the exact role of H3K27me3 in hypertension remains unknown [21–26, 28]. Increased H3K27me3 levels have been associated with changes in blood pressure [53]. However, the exact genes containing H3K27me3 alterations have yet to be identified. We identify that H3K27me3 enrichment at *EDNRB*, *ACTA2*, *TAGLN*, *CNN1*, and *MYH11* gene promoters is regulated by JMJD3, and future studies will determine how H3K27me3 at these promoters changes in the setting of hypertension. Our identification of *EDNRB* as a direct transcriptional target of JMJD3 is consistent with reports that have identified that *EDNRB* expression is positively regulated by transcription factors (e.g., MKL2) and mechanisms that regulate SMC-specific gene expression [54]. Upon disease progression, decreased JMJD3 expression further leads to repression of the SMC gene program (via increased endothelin-ERK signaling and H3K27me3 at SMC promoters), thereby promoting vascular remodeling in the setting of long-standing hypertension. While SMCs are the predominant cell type regulating vessel tone and blood pressure, endothelin signaling involves interplay between SMCs and endothelial cells (ECs). Therefore, it is possible that JMJD3 in both SMCs and ECs synergistically regulates endothelin-dependent blood pressure to result in a more severe disease phenotype. This scenario may be addressed with more highly powered GTEx data for rs62059712 in SMCs and ECs, since current GTEx data is largely inconclusive, most likely due to the relatively low frequency of the rs62059712 minor allele.

We demonstrate that the rs62059712 major T allele decreases *JMJD3* expression, mechanistically resulting in a “double hit,” first leading to increased blood pressure via increased *EDNRA* expression, and next, resulting in increased pathologic remodeling due to increased ERK signaling and H3K27me3 at smooth muscle gene promoters. Our findings support a translational role for rs62059712. In particular, we propose that rs62059712 genotype can be used to identify patients who may be more likely to respond to endothelin receptor antagonists, which are currently used for pulmonary artery hypertension and in clinical trails for the treatment of essential hypertension. Furthermore, targeting the endothelin and/or ERK pathway(s) in certain patients may also help reduce arterial remodeling due to long-standing hypertension.

Blood pressure is a complex phenotype controlled by multiple factors. Our study identifies the role of rs62059712 in its association with blood pressure. We show that the major T allele decreases *JMJD3* transcription in SMCs by disrupting SP1 binding to the *JMJD3* promoter, leading to decreased *JMJD3* expression. We identify the *EDNRB* gene as a direct transcriptional target of JMJD3, which is decreased in the setting of the major T allele, thereby resulting in compensatory increase in *EDNRA* expression. Thus, loss of *JMJD3* increases endothelin signaling and downstream vessel contractility, which is reduced by treatment with the dual endothelin A and B antagonist, bosentan. Decreased *JMJD3* further negatively modifies disease phenotype by leading to increased ERK activation, which increases SMC migration and phenotypic modulation, two critical events that occur during pathologic arterial remodeling. In conclusion, our findings define a novel transcriptional and molecular axis involving the histone demethylase, JMJD3, in SMCs that regulates blood pressure and arterial response to injury.

## Data Availability

All data in the manuscript will be made publicly available upon publication of the manuscript.

## Acknowledgements

We thank Robin G. Kunkel, research associate in the Pathology Department, University of Michigan, for her artistic work.

## Author contributions

Conceptualization: KDM, KAG

Experimental Design: KDM, LC, AO, FMD, LCT, JG, KAG

Performed experiments: KDM, LQ, TB, SJW, JS, JYM, ECB, ADJ, ZA, RW, KB

Data analysis: KDM, LQ, ADJ, RW, LCT

Figures: KDM, LQ Writing: KDM, KAG

Funding acquisition: KDM, KAG

